# Insulin levels early in perimenopause inform vasomotor symptom incidence across the menopausal transition

**DOI:** 10.1101/2025.09.05.25334479

**Authors:** Faria Athar, Sarah Gregory, Emma J. Houston, Nicole M. Templeman

## Abstract

**Context:** Metabolic health and body mass index (BMI) has been implicated in influencing menopause-related changes, but the role of elevated insulin, an early marker of metabolic dysfunction, remains understudied.

**Objective:** To determine whether midlife fasting insulin is associated with menopausal changes such as vasomotor symptom incidence and reproductive hormone trajectories.

**Methods:** This study consisted of longitudinal analyses of 704 community-based participants from the Study of Women’s Health Across the Nation (SWAN). It included data from baseline to the 10th annual visit, restricted to women who had age-47 fasting insulin measurements. Main outcome measures included timing and duration of hot flashes, night sweats, cold sweats, and vaginal dryness, and trajectories of estradiol, follicle-stimulating hormone, and testosterone from 6 years before to 6 years after the final menstrual period.

**Results:** Higher fasting insulin levels during early perimenopause predicted earlier onset of hot flashes and night sweats, longer duration of hot flashes and cold sweats, and greater increases in testosterone. In Cox proportional hazards models, elevated age-47 insulin was associated with a higher hazard of hot flashes and cold sweats; the association between insulin and hot flash likelihood remained significant when BMI was incorporated as a covariate. BMI associations with vasomotor symptoms paralleled those of insulin levels, but BMI appeared to be more closely associated with slower estradiol decline and blunted FSH rise.

**Conclusions:** Perimenopausal fasting insulin and BMI show complementary but distinct associations with menopausal changes. Elevated insulin predicts earlier and prolonged vasomotor symptoms, and is also associated with higher testosterone levels.

## Introduction

The menopausal transition is accompanied by physiological changes that diminish quality of life. These may include hot flashes, night sweats, or cold sweats, which are often collectively described as vasomotor symptoms, as well as other associated changes such as vaginal dryness, changes in blood pressure, and heart palpitations (Talaulikar, 2022). Vasomotor symptoms affect approximately three-quarters of women during the menopausal transition (Gold et al., 2006; Woods and Mitchell, 2005); they can appear around 2 years before the final menstrual period, and persist for as long as 8-10 years beyond it (Avis et al., 2018). The severity and duration of these symptoms can be affected by factors such as race, ethnicity, and socioeconomic status. For instance, Black women report having a high prevalence and long duration of vasomotor symptoms, while women of Asian descent report low vasomotor symptom prevalence (Blanken et al., 2022; Lee et al., 2010). However, women of the same ethnicity may also have varying degrees of symptoms based on their location and socioeconomic status (Gold et al., 2006; Gupta et al., 2006; Tepper et al., 2016). Vasomotor symptoms and other physical features of the menopausal transition contribute to sleep disruption, fatigue, anxiety, and depression, which leads to repercussions for social relationships and overall wellbeing (Nappi et al., 2021). While hormone replacement therapy serves as the primary approach to alleviate vasomotor symptoms, concerns about long-term risks and inequities in availability (Conklin et al., 2024; Iyer and Manson, 2024; Khan et al., 2023; Palacios et al., 2019) has prompted exploration into alternative treatment and management strategies to delay the onset and mitigate the severity of vasomotor symptoms.

Although our understanding of vasomotor symptom pathophysiology is limited, a primary driver appears to be estrogen withdrawal and the related hypothalamic shift that accompanies perimenopause (Deecher and Dorries, 2007; Rapkin, 2007). Rising levels of follicle stimulating hormone (FSH) precede the decline in estrogen levels, with FSH rising approximately a year before estrogen reaches its lowest point (Hall, 2015). Testosterone (T) levels might remain relatively stable or decrease around the menopausal transition (Burger, 2006). Impacts of these endocrine shifts extend beyond the hypothalamus-pituitary-ovary axis. For instance, overactivation of the KNDy neurons due to low estrogen levels can create a narrowed thermoneutral zone, making women more susceptible to temperature fluctuations that trigger the flushing, sweating, and peripheral vasodilation that are characteristic of hot flashes (Gombert-Labedens et al., 2025; Meczekalski et al., 2025; Padilla et al., 2018). Estrogen withdrawal from other tissues induces other changes, such as reducing tissue flexibility and elasticity, which can cause thinner and drier vaginal walls and the resultant sexual dysfunction (Mac Bride et al., 2010; Palma et al., 2016).

Recently, metabolic health has emerged as an important but relatively underexplored determinant of menopausal changes. For instance, women with obesity and higher body fat percentages report having an increased frequency and severity of vasomotor symptoms (Namgoung et al., 2022; Tang et al., 2022) and an increased incidence of vasomotor symptoms early in menopause (Gold et al., 2017). Insulin resistance and high HbA1C levels, both markers of poor glycemic control, are also associated with a greater disposition to vasomotor symptoms (Kwon et al., 2016; Rouen et al., 2015; Thurston et al., 2012). Increased vasomotor symptom severity and/or frequency has also been associated with a greater risk of a subsequent type 2 diabetes diagnosis (Gray et al., 2018; Hedderson et al., 2024). Since rates of obesity and metabolic dysfunction have been rising globally, there is an increased likelihood that more individuals worldwide will experience difficult menopausal transitions and poorer subsequent health outcomes (Thurston and Joffe, 2011).

A wide array of physiological changes can accompany or exacerbate deteriorations in metabolic health, but elevated insulin levels have emerged as an early feature—and potentially, an upstream driver—of metabolic disorders such as obesity, insulin resistance, and type 2 diabetes (Johnson, 2021; Thomas et al., 2019; Tricò et al., 2018; Zhang et al., 2021). Although racial differences exist in insulin dynamics (Chung et al., 2019), hyperinsulinemia is generally defined as insulin concentrations that exceed the normal range relative to an individual’s age and health status, without accompanying hypoglycemia (Thomas et al., 2019). Insulin is best known for its role in glucose homeostasis, but it also has important actions on the female reproductive system, as well as intricate relationships with sex steroid signaling (Athar et al., 2024). Insulin receptors are distributed throughout the central nervous system, including GnRH neurons, astrocytes, and Kiss1 neurons, and they are also expressed on oocytes and somatic cells of the ovary (Acevedo et al., 2007; Dupont and Scaramuzzi, 2016). Through multiple sites of action, insulin signalling regulates reproductive function by such means as modulating GnRH pulsatility and gonadotropin secretion, and it can also act synergistically as a co-gonadotropin to augment ovarian steroidogenesis (Evans et al., 2021; Sliwowska et al., 2014). Given that insulin plays a role in directing reproductive processes and that its levels may be elevated early in the development of metabolic disorders, we wished to investigate whether insulin levels prior to menopause might predict the severity and progression of physical menopause symptoms.

In our analyses, we used data from the Study of Women’s Health Across the Nation (SWAN) that longitudinally followed women from a pre-/perimenopausal baseline (42-52 years of age) across the subsequent 10 years (Sowers et al., 2000). To evaluate metabolic health at an early study time point that had an adequate sample size, we assessed metabolic measurements at age 47, and related these to physiological features of the menopausal transition.

## Materials and methods

These secondary analyses used anonymized, publicly available data from SWAN, including biological, behavioral, and socio-economic data from baseline up to the 10th follow-up visit, all of which were collected between 1995 and 2008. The Study of Women’s Health Across the Nation was a multi-site longitudinal cohort study of middle-aged women across seven sites in the United States. The detailed study design and protocol have been described previously (Sowers et al., 2000). The SWAN study protocol was approved by the institutional board at each study site, and all participants provided written informed consent. Our secondary data analyses were approved by the Human Research Ethics Board at the University of Victoria (protocol number: 25-0295).

The 3302 participants who were enrolled in SWAN at baseline had an intact uterus and one or more ovaries, were not taking hormone therapy, not pregnant or lactating and had experienced one or more menses in the prior 3 months. Here, we restricted our analyses to women who also satisfied the following criteria: they attended a study visit at the age of 47, and they had complete data for insulin measurements at that visit. Participants were excluded from our analyses if they were taking insulin medications to manage blood sugar or other health conditions.

The measurements taken in the SWAN protocol have previously been described in detail (Sowers et al., 2000). In brief, blood was collected after an overnight fast and centrifuged to collect serum for analysis. Insulin was measured in serum by a solid phase radioimmunoassay. Fasting insulin values from study participants when they were 47 years old were log-transformed to normalize the distribution for our analyses. Estradiol (E2) levels were assayed using a double-antibody chemiluminescent immunoassay. FSH was measured using a two site chemiluminometric immunoassay. The total testosterone (T) assay was a modified rabbit polyclonal anti-T immunoassay. Body mass index (BMI) was measured at the annual clinic visits using a stadiometer and balance beam and was determined as weight in kg divided by height^2^ in meters.

Participants self-reported the occurrence of hot flashes, night sweats, cold sweats, and vaginal dryness in the two weeks prior to the visit. We defined the duration of experiencing a symptom across the 10-year study period as the number of annual visits over which a symptom was reported, and we also calculated the age of onset of each symptom. Covariates included self-reported race/ethnicity, smoking status, and income at the baseline visit. Final menstrual period was estimated as the date of last menstrual period preceding 12 months of amenorrhea.

Linear regression models were used to examine associations between log-transformed insulin levels or BMI at age 47 and continuous symptom measures. We first fit unadjusted simple linear regression models, followed by multiple linear regression models adjusted for self-reported race/ethnicity, income, and smoking status to account for differences in both insulin levels and menopausal symptom patterns. Ordinary least squares estimation was applied, with statistical significance set at p < 0.05. Hormone trajectory slopes relative to final menstrual period were estimated for each participant using mixed-effects models with log-transformed hormone values, and these individual slopes were then regressed on 47-year-old BMI and log-insulin levels using linear regression, adjusting for demographic covariates. To further examine the association between insulin or BMI and symptom incidence, we used Cox proportional hazards models. For each symptom, we fitted sequential models examining insulin alone, BMI alone, both predictors simultaneously, and insulin × BMI interactions, with hazard ratios representing risk per one standard deviation increase in predictor. We did complete case analysis and analyses were conducted in RStudio (R version 4.1.1).

## Results

Our final dataset included 704 participants. The self-reported racial and ethnic composition of the included participants was: 375 (53.3%) Caucasian/White Non-Hispanic, 149 (21.2%) Black/African American, 98 (13.9%) Japanese/Japanese American, and 82 (11.6%) Chinese/Chinese American. Mean insulin levels were 10.117 µIU/mL (SD = 6.711 µIU/mL), and BMI had a mean of 27 kg/m^2^ (SD = 6.6 kg/m^2^). At the age of 47, a majority of women in our dataset reported an absence of physical menopausal symptoms (69%, 69.1%, 90.9%, and 78.3% reported that they did not experience hot flashes, night sweats, cold sweats, and vaginal dryness, respectively) over the preceding 2-week period. The average age of final menstrual period for the individuals included in our dataset was 51 years.

### Insulin levels predict age of onset and duration of symptoms of the menopausal transition

Five symptom variables showed statistically significant associations with insulin levels (Table 1). The most pronounced unadjusted associations included the ages of onset for hot flashes (β = -1.54, p < 0.001), night sweats (β = -0.947, p = 0.003), and vaginal dryness (β = -0.774, p = 0.039), all associations which indicated earlier symptom onset if the individuals had higher fasted insulin levels at 47 years of age. In addition, higher insulin was associated with experiencing longer durations of cold sweats (β = 0.652, p < 0.001) and/or hot flashes (β = 0.959, p < 0.001). The durations of experiencing night sweats and vaginal dryness, however, were not significantly associated with insulin levels (β = 0.288, p = 0.306; and β = 0.043, p = 0.879; respectively). Four of these five significant relationships remained significant in models adjusted for self-reported race, income, and smoking status. These included the ages of onset of hot flashes (β = -1.14, p < 0.001) and night sweats (β = -0.69, p = 0.035), and the durations of hot flashes (β = 0.62, p = 0.032) and cold sweats (β = 0.38, p = 0.018). The age of onset of vaginal dryness lost statistical significance in the adjusted model (β = -0.66, p = 0.095). Collectively, these data indicate that women with higher insulin levels at 47 years of age have an increased likelihood of experiencing vasomotor symptoms earlier, and for a longer duration of time.

**Table 1.** Crude and adjusted associations between age-47 fasting insulin levels and menopause symptoms.

| Exposure: Insulin<br>Menopause Symptom | Unadjusted |  |  | Adjusted for self-reported race + income + smoking status |  |  |
| --- | --- | --- | --- | --- | --- | --- |
|  | Coeff | p-value | Adj R<sup>2</sup> | Coeff | p-value | Adj R<sup>2</sup> |
| **HOT FLASHES** |  |  |  |  |  |  |
| Age of Onset | **-1.543** | **< 0.001** | **0.044** | **-1.138** | **< 0.001** | **0.077** |
| Duration | **0.959** | **< 0.001** | **0.017** | **0.618** | **0.032** | **0.064** |
| **NIGHT SWEATS** |  |  |  |  |  |  |
| Age of Onset | **-0.947** | **0.003** | **0.017** | **-0.692** | **0.035** | **0.044** |
| Duration | 0.288 | 0.306 | 0.001 | -0.087 | 0.759 | 0.09 |
| **COLD SWEATS** |  |  |  |  |  |  |
| Age of Onset | -0.855 | 0.074 | 0.013 | -0.593 | 0.226 | 0.075 |
| Duration | **0.652** | **< 0.001** | **0.022** | **0.379** | **0.018** | **0.125** |
| **VAGINAL DRYNESS** |  |  |  |  |  |  |
| Age of Onset | **-0.774** | **0.039** | **0.009** | -0.663 | 0.095 | 0.014 |
| Duration | 0.043 | 0.879 | <0.001 | -0.057 | 0.85 | <0.001 |

### Higher insulin and a higher BMI at age 47 are parallel predictors of vasomotor symptoms

BMI is a widely used metric to approximate physical fitness based on the height and weight ratios. A higher BMI is associated with worsened outcomes of key changes during the menopausal transition (Opoku et al., 2023), but BMI is an imprecise metric that does not account for muscle mass, adipose distribution, or ethnic differences in body composition. Thus, we wished to assess how insulin levels compare to BMI in terms of predicting physical menopause symptoms. In simple linear regressions without adjustment, we observed that the ages of onset of all tested symptoms were significantly associated with a higher BMI at age 47. Additionally, the duration of hot flashes (β = 0.0799, p < 0.0001), night sweats (β = 0.0628, p = 0.0009), and cold sweats (β = 0.0597, p < 0.0001) was also significantly associated with higher BMI. After accounting for self-reported race and socioeconomic factors, three relationships maintained their statistical significance: the ages of onset of hot flashes (β = -0.0931, p < 0.001) and night sweats (β = -0.0469, p = 0.045), and the duration of experiencing cold sweats (β = 0.0246, p = 0.031; Table 2).

**Table 2.** Crude and adjusted associations between age-47 BMI and menopause symptoms.

| Exposure: BMI | Unadjusted |  |  | Adjusted for self-reported race + income + smoking status |  |  |
| --- | --- | --- | --- | --- | --- | --- |
| Menopause Symptom | Coeff | p-value | Adj R<sup>2</sup> | Coeff | p-value | Adj R<sup>2</sup> |
| **HOT FLASHES** |  |  |  |  |  |  |
| Age of Onset | **-0.1215** | **< 0.0001** | **0.059** | **-0.0931** | **< 0.0001** | **0.083** |
| Duration | **0.0799** | **< 0.0001** | **0.025** | 0.0375 | 0.069 | 0.062 |
| **NIGHT SWEATS** |  |  |  |  |  |  |
| Age of Onset | **-0.0744** | **0.0004** | **0.022** | **-0.0469** | **0.045** | **0.043** |
| Duration | **0.0628** | **0.0009** | **0.016** | 0.0042 | 0.835 | 0.09 |
| **COLD SWEATS** |  |  |  |  |  |  |
| Age of Onset | **-0.0883** | **0.007** | **0.029** | -0.0533 | 0.136 | 0.078 |
| Duration | **0.0597** | **< 0.0001** | **0.041** | **0.0246** | **0.031** | **0.124** |
| **VAGINAL DRYNESS** |  |  |  |  |  |  |
| Age of Onset | **-0.057** | **0.021** | **0.012** | -0.0385 | 0.165 | 0.012 |
| Duration | -0.0015 | 0.939 | <0.001 | -0.0026 | 0.905 | <0.001 |

In our analyses, both insulin levels and BMI emerged as important predictors of menopause symptom timing and duration. Adjustments for sociodemographic factors caused a higher proportion of the BMI relationships to lose statistical significance, suggesting that BMI may be more confounded by factors such as race/ethnicity than insulin levels. The close tracking between adjusted R^2^ values for insulin and BMI (Figure 1) raises the possibility that insulin dysregulation, rather than obesity alone, may play a key role in mediating BMI-associated changes in vasomotor symptom burden during menopause.

**Figure 1.**
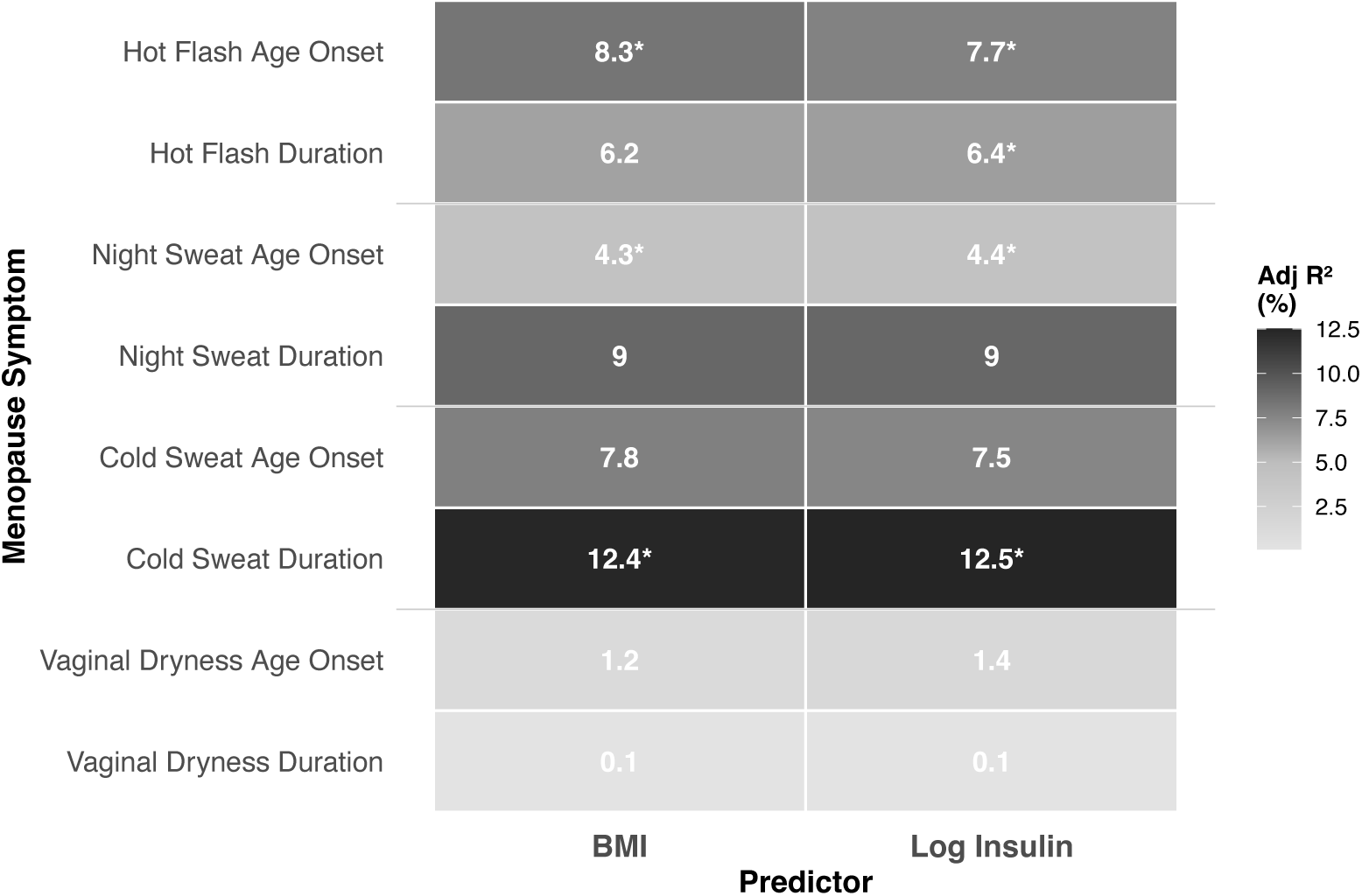
Parallels between effects of age-47 BMI and insulin on menopause symptoms. Adjusted R² values (%) for associations between age-47 BMI and log insulin with menopause symptom onset and duration. Heat map intensity reflects association strength. * indicates p < 0.05.

To use a different approach for comparing the predictive capacities of these two metabolic features, we performed Cox proportional hazards regressions with BMI and fasting insulin values at the age of 47 (Table 3). After adjusting for self-reported race/ethnicity, income, and smoking status, insulin levels at age 47 showed stronger predictive effects for incidence of key menopause symptoms. For instance, each standard deviation rise in log-insulin (*i.e.*, an increase of 5.6 μIU/ml in fasting insulin levels) was associated with a 14% higher hazard of hot flashes (HR = 1.14, 95% CI 1.05-1.24, p = 0.002), while BMI did not show a statistically significant effect (HR = 1.07, 95% CI 0.99-1.17, p = 0.104). Higher insulin levels also predicted a 20% higher hazard of cold sweats (HR = 1.20, 95% CI 1.06-1.35, p = 0.004), compared to an 18% increase for BMI (HR = 1.18, 95% CI 1.04-1.34, p = 0.012). In multivariable models that included both predictors (*i.e.*, both insulin and BMI), age-47 fasting insulin levels maintained independent significance for a positive association with hot flash likelihood (HR = 1.15, 95% CI 1.04-1.27, p = 0.007) while none of the BMI effects reached the threshold of statistical significance once insulin was incorporated as a covariate. Therefore, fasting insulin levels may have more pronounced effects on the instantaneous risk of vasomotor symptoms. Elevated insulin levels, which can be early indicators of metabolic dysfunction, may serve as valuable markers for identifying individuals at risk for early and potentially severe vasomotor symptoms.

**Table 3.** Univariate and multivariate Cox regression analyses of age-47 insulin and BMI effects on menopausal symptom onset.

| Symptom | Univariate model |  |  |  | Multivariate model |  |  |  |
| --- | --- | --- | --- | --- | --- | --- | --- | --- |
|  | Insulin HR<br>(95% CI) | p-value | BMI HR<br>(95% CI) | p-value | Insulin HR<br>(95% CI) | p-value | BMI HR<br>(95% CI) | p-value |
| Hot<br>Flashes | **1.14**<br>**(1.05-1.24)** | **0.002** | 1.07<br>(0.99-1.17) | 0.104 | **1.15**<br>**(1.04-1.27)** | **0.007** | 0.99<br>(0.90-1.10) | 0.894 |
| Night<br>Sweats | 1.05<br>(0.96-1.14) | 0.321 | 1.06<br>(0.96-1.16) | 0.258 | 1.03<br>(0.93-1.14) | 0.632 | 1.04<br>(0.93-1.16) | 0.472 |
| Cold<br>Sweats | **1.20**<br>**(1.06-1.35)** | **0.004** | **1.18**<br>**(1.04-1.34)** | **0.012** | 1.14<br>(0.99-1.32) | 0.063 | 1.10<br>(0.94-1.27) | 0.234 |
| Vaginal<br>Dryness | 1.01<br>(0.91-1.11) | 0.905 | 1.00<br>(0.90-1.12) | 0.964 | 1.01<br>(0.90-1.13) | 0.91 | 1.00<br>(0.88-1.14) | 0.982 |

### BMI is linked to FSH and estrogen dynamics while insulin is associated with testosterone

Vasomotor symptoms like hot flashes are also tied to fluctuations in estrogen and FSH (Randolph et al., 2005). Thus, we wished to understand how BMI and insulin levels inform changes in levels of these hormones across the menopausal transition. In our analyzed dataset, we observed menopausal hormonal changes that were largely consistent with prior reports: estradiol levels decreased across the menopausal transition (-64.58%), while FSH levels rose markedly by 564.37% (Figure 2). However, our dataset showed a slight, 6.78% average rise in testosterone levels over the same window of six years preceding and six years following the final menstrual period. We found that overall, a higher BMI at age 47 was associated with a blunted decline in estradiol and a more-gradual rise in FSH levels across the menopausal transition. In contrast, insulin levels were more closely associated with testosterone changes, with higher insulin linked to a greater rise in testosterone over time (Table 4; Figure 2). These data suggest that while BMI may shape the trajectories of gonadotropins and estrogens during the menopausal transition, insulin may be more related to androgen-related hormone changes.

**Figure 2.**
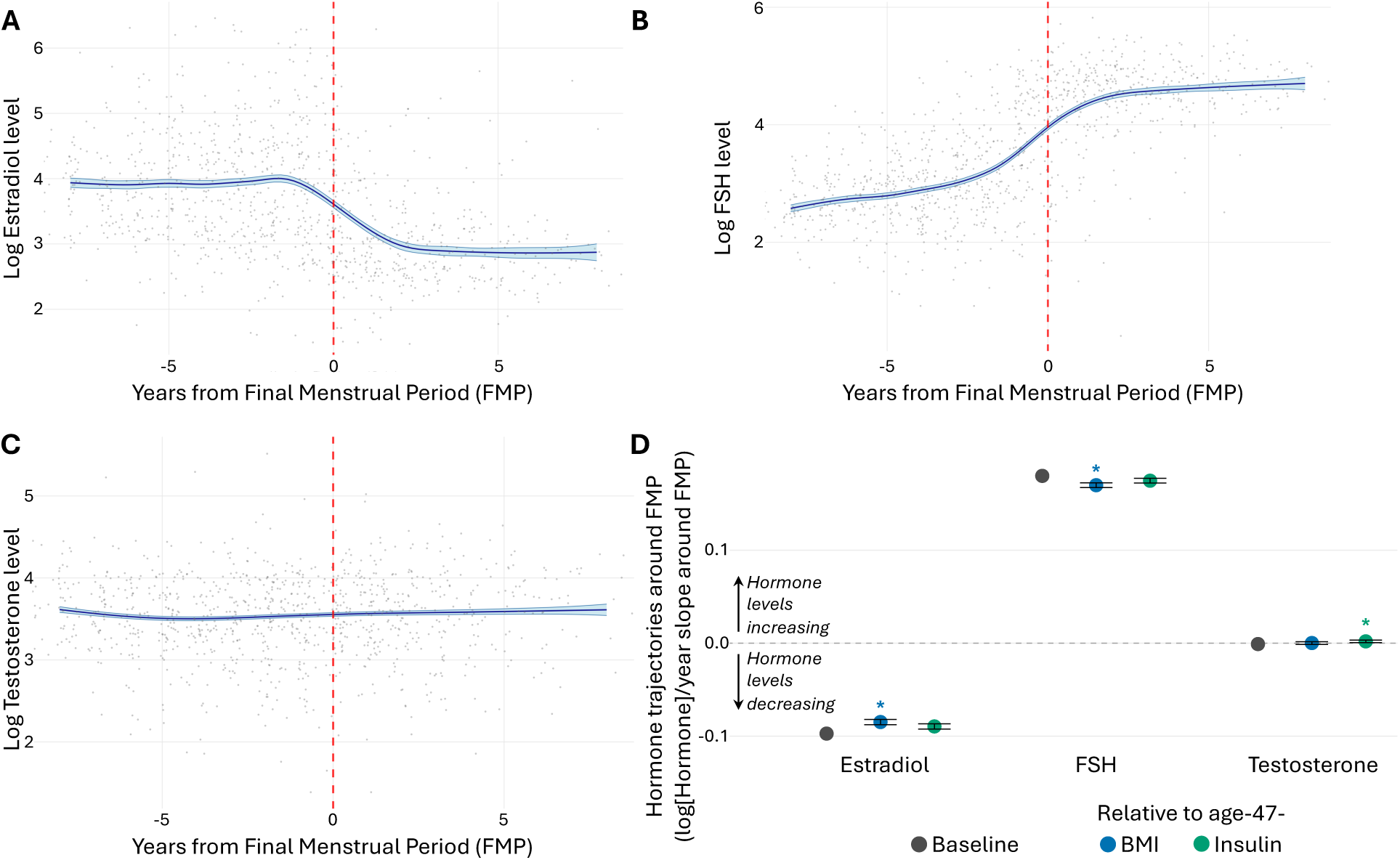
Hormone trajectories during the menopausal transition, and associations with age-47 insulin and BMI. Panels A-C show trajectories of log-transformed hormone levels relative to the final menstrual period (FMP, indicated by red dashed line). (A) Estradiol levels decline sharply around the FMP. (B) FSH levels rise progressively, with peak occurring after the FMP. (C) Testosterone levels remain relatively stable throughout the transition. Gray points represent individual observations; blue lines show fitted curves with 95% confidence intervals. (D) displays slopes (or trajectories of hormone level changes around the FMP) for estradiol, FSH and testosterone in relation to BMI (blue) and insulin (green); baseline change in slopes are represented in gray. Direction of change from FMP indicated as arrows from point 0.0, with positive values denoting an increase in hormone levels from 6 years prior to 6 years post-FMP, and negative values indicating decreasing hormone levels in that time period. Significant associations are denoted by * and bars represent 95% confidence intervals. N = 690.

**Table 4.** Associations of fasting insulin and BMI at age 47 with hormone trajectories across the menopausal transition.

| Hormone Trajectory | BMI Coefficient | BMI p-value | BMI Adj R<sup>2</sup> | Insulin Coefficient | Insulin p-value | Insulin Adj R<sup>2</sup> |
| --- | --- | --- | --- | --- | --- | --- |
| Estradiol | **0.0017** | **<0.001** | **0.109** | 0.0039 | 0.291 | 0.052 |
| FSH | **-0.0015** | **<0.001** | **0.078** | 0.0001 | 0.971 | 0.022 |
| Testosterone | -0.0001 | 0.335 | 0.022 | **0.0074** | **<0.001** | **0.043** |

## Discussion

In this study, we used longitudinal data from the SWAN study to examine how insulin levels and BMI influence physical changes of menopause. We found that insulin and BMI affect the menopausal transition in distinct but complementary ways. These two metabolic features aligned in their overall impacts on the timing and duration of vasomotor symptoms, yet pre-/peri-menopausal fasting insulin may be especially effective at predicting incidence of these symptoms, particularly hot flashes. Meanwhile, BMI is more closely associated with the trajectories of the reproductive hormones FSH and estradiol. These data together suggest that metabolic health at the onset of perimenopause can cumulatively inform many aspects of the menopause experience.

Higher levels of fasting insulin at age 47 were associated with an earlier and increased risk of vasomotor symptoms. In parallel to these relationships with insulin levels, we also observed that BMI at 47 is associated with earlier and extended experience of vasomotor symptoms. This aligns with prior work indicating that BMI and waist circumference have greater associations with vasomotor symptom onset earlier rather than later in menopause (Gold et al., 2017). Since insulin continued to significantly predict the likelihood of experiencing hot flashes in combined Cox regression models, independent of confounding effects of BMI, our analyses point to the relevance of considering pre/peri-menopausal insulin levels as a metric for better understanding and forecasting menopause symptoms. Interestingly, rodent studies have shown that insulin may have hyperthermic effects through its actions on astrocytes and hypothalamic neurons that regulate thermogenesis (Manaserh et al., 2020; Sanchez-Alavez et al., 2009).

We observed distinct associations between insulin versus BMI and the trajectories of sex hormone changes across the menopausal transition. Higher BMI was linked to a slower decline in estradiol and a slower rise in FSH. This is consistent with prior research showing that adiposity may influence estrogen and FSH feedback, such that obese post-menopausal women have higher estradiol and lower FSH than their lean counterparts, likely due to peripheral effects of obesity on gonadotropins (Freeman et al., 2010; Kim et al., 2015; Liu et al., 2022). In contrast, insulin levels were more strongly associated with a steeper increase in testosterone levels around menopause. This pattern is characteristic of a hyperandrogenic state, which are usually associated with hyperinsulinemia as well as with poor cardiovascular outcomes (Ichikawa et al., 2025; Stangl et al., 2024; Sutton-Tyrrell et al., 2005; Weinberg et al., 2006). Higher postmenopausal levels of testosterone and/or estradiol have also been prospectively related to increased risk of type 2 diabetes (Ding et al., 2007, 2006; Muka et al., 2016). Together, these data suggest that BMI is associated with the dynamics of estrogen withdrawal and gonadotropin response, while insulin appears more tightly linked to androgen profiles.

Importantly, insulin levels can be modulated. Reversing obesity is dependent on a multitude of factors and can be especially challenging among some ethnicities and socio-economic groups (Hill-Briggs et al., 2020; Pinchevsky et al., 2020). In fact, women tend to experience greater difficulty losing weight than men (Li et al., 2020). Conversely, insulin levels may be more sensitive than body weight to lifestyle interventions like exercise and a healthy diet (Bird and Hawley, 2017; Brown et al., 2020). Aerobic and resistance exercise can lower insulin levels and improve insulin sensitivity independent of body composition and weight loss (Kim and Park, 2013). Given that reductions in insulin levels often precede intervention-induced weight loss (Wiebe et al., 2021), improving metabolic health through mitigating high insulin levels may offer a more-attainable target.

Our findings align with previous work linking insulin resistance with worsened menopause symptoms. Insulin resistance is defined as the diminished capacity for insulin-stimulated glucose disposal, and it has a close interrelationship with hyperinsulinemia (Johnson, 2021; Thomas et al., 2019). Higher values for HOMA-IR, a surrogate index of insulin resistance calculated from fasting glucose and insulin levels, have been previously associated with hot flashes and night sweats, with especially strong associations reported for hot flashes (Thurston et al., 2012); our findings suggest that elevated fasting insulin itself may be a key component of this relationship. While insulin resistance and hyperinsulinemia frequently co-occur, emerging evidence suggests that hyperinsulinemia can be detected in the absence of insulin resistance, that it may precede insulin resistance and obesity, and that it may even contribute to their development (Johnson, 2021; Thomas et al., 2019; Tricò et al., 2018). Thus, the distinction between elevated insulin and insulin resistance might be especially relevant for midlife women who could be experiencing early shifts in metabolic health with potential repercussions for hormone dynamics and symptom incidence across the menopausal transition.

## Study limitations

Although the SWAN study has the strengths of longitudinally tracking women from a large and diverse cohort, insulin and other hormone levels were measured annually and do not capture the full extent of hormone fluctuations. The participants who had relatively higher fasting insulin levels may or may not have also been insulin resistant; based on available data, we could not preclude the possibility that insulin resistance plays a role in the relationships we detected, although we did observe that elevated fasting insulin was itself associated with specific hormone and symptom patterns. Only fasting insulin levels were included in these data, which did not allow us to assess postprandial or dynamic insulin responses. Prior work has shown that being climacteric at 46 years of age is independently associated with elevated insulin levels after a glucose challenge (Savukoski et al., 2021), which highlights the possibility that dynamic metabolic assessments may reveal further relationships with menopausal symptoms. By design, the SWAN study recruited women who were likely to be already perimenopausal, which limited our ability to establish predictive relationships earlier in life. Notably, women undergoing an early menopause also show differences in insulin profiles (Roa-Díaz et al., 2023). Nevertheless, our analyses suggest that insulin and BMI early in perimenopause exert distinct effects on reproductive hormone trajectories and symptom timing during the menopausal transition. Insulin may be a stronger predictor than BMI for certain outcomes, particularly androgenic hormone changes and vasomotor symptom incidence. These insights could improve understanding of how premenopausal or perimenopausal metabolic health shapes the experience and long-term repercussions of the menopausal transition.

## Data Availability

All data produced in the present work are contained in the manuscript.

## Acknowledgements

We thank the study staff at each site and all women who participated in SWAN. We also appreciated guidance and insights of Graciela Muniz-Terrera and Andrea Piccinin for these analyses.

Clinical Centers: University of Michigan, Ann Arbor – Carrie Karvonen-Gutierrez, PI 2021–present, Siobán Harlow, PI 2011–2021, MaryFran Sowers, PI 1994–2011; Massachusetts General Hospital, Boston, MA – Sherri-Ann Burnett-Bowie, PI 2020–Present; Joel Finkelstein, PI 1999–2020; Robert Neer, PI 1994–1999; Rush University, Rush University Medical Center, Chicago, IL – Imke Janssen, PI 2020– Present; Howard Kravitz, PI 2009–2020; Lynda Powell, PI 1994–2009; University of California, Davis/Kaiser – Elaine Waetjen and Monique Hedderson, PIs 2020–Present; Ellen Gold, PI 1994–2020; University of California, Los Angeles – Arun Karlamangla, PI 2020 – Present; Gail Greendale, PI 1994– 2020; Albert Einstein College of Medicine, Bronx, NY – Carol Derby, PI 2011–present, Rachel Wildman, PI 2010–2011; Nanette Santoro, PI 2004–2010; University of Medicine and Dentistry – New Jersey Medical School, Newark – Gerson Weiss, PI 1994–2004; and the University of Pittsburgh, Pittsburgh, PA – Rebecca Thurston, PI 2020–Present; Karen Matthews, PI 1994–2020. Steering Committee: Susan Johnson, Current Chair. Chris Gallagher, Former Chair.

## Notes

Funding: Templeman Lab research is supported by funding from the Canadian Institutes of Health Research (PJT - 183618). N.M.T. is a Tier 2 Canada Research Chair in Cell Biology, a Michael Smith Health Research BC Scholar, and a member of the Sexual and Reproductive Health and Rights Research Cluster at the University of Victoria.

### Competing Interest Statement

The authors have declared no competing interest.

### Funding Statement

Templeman Lab research is supported by funding from the Canadian Institutes of Health Research. N.M.T. is a Tier 2 Canada Research Chair in Cell Biology, a Michael Smith Health Research BC Scholar, and a member of the Sexual and Reproductive Health and Rights Research Cluster at the University of Victoria.
The Study of Womens Health Across the Nation (SWAN) has grant support from the National Institutes of Health (NIH), Department of Health and Human Services (DHHS), through the National Institute on Aging (NIA), the National Institute of Nursing Research (NINR) and the NIH Office of Research on Womens Health (ORWH).

### Author Declarations

The Human Research Ethics Board of the University of Victoria gave ethical approval for this work.

